# Understanding, Confidence and Usefulness of Effect Size and Certainty of Evidence: A Cross-Sectional Survey Among Australian Dietitians

**DOI:** 10.64898/2026.11.03.26366416

**Authors:** Nirjhar R. Ghosh, Elizabeth P. Neale, Bradley C. Johnston

**Affiliations:** Department of Nutrition, College of Agriculture and Life Sciences, Texas A&M University, College Station, Texas, USA; Department of Surgery, Houston Methodist Hospital, Houston, TX, USA; School of Medical, Indigenous and Health Sciences, Faculty of Science, Medicine and Health, University of Wollongong, Wollongong, NSW, Australia; Department of Nutrition, College of Agriculture and Life Sciences, Texas A&M University, College Station, USA; Department of Epidemiology & Biostatistics, School of Public Health, Texas A&M University, College Station, TX, USA

**Keywords:** clinical nutrition, evidence-based dietetic practice, evidence-based practice competencies, treatment effect size, certainty of evidence

## Abstract

**Purpose:** Interpreting treatment effect size is essential for evidence-sinformed dietetic care, yet dietitians’ understanding remains unestablished.

**Methods:** We conducted a web-based cross-sectional survey of Australian dietitians (October 2024-February 2025). The survey assessed their understanding of effect size, presented using three absolute estimates (risk difference [RD], risk in treatment vs. risk in control groups, number needed to treat [NNT]) and two relative estimates (relative risk [RR], odds ratio [OR]). We also assessed dietitians’ confidence in determining correct effect size, their perceived usefulness of effect estimates and corresponding certainty of evidence.

**Results:** Approximately 7,697 dietitians received the survey link; 27 participated and 13 completers were analyzed. Correct understanding was higher for absolute than relative estimates (30.0% vs. 18.2%). Risk in treatment vs. risk in control groups was understood the most (41.7%) and RD, RR and OR the least (18.2% each). Confidence was highest for RR [mean (SD): 4.18 (1.25)] and lowest for NNT [3.40 (1.17)]. Perceived usefulness was highest for OR [4.69 (1.18)] and lowest for NNT [3.75 (1.76)]. Certainty of evidence was rated useful [5.69 (1.03)].

**Conclusion:** Based on a small sample, preliminary evidence suggests limited understanding of effect size, highlighting the need for further training and assessment in larger sample.

## INTRODUCTION

High-quality dietetic care depends on integrating the best available evidence with clinical expertise and patients’ values and preferences (1-3), an approach endorsed by Dietitians Australia’s National Competency Standards (4). Because up-to-date systematic reviews with meta-analyses (SRMAs) are considered best evidence for clinical decision-making, dietitians are encouraged to use SRMA findings in practice (5-7). These findings are presented as summary estimates that describe treatment effect size, and for dichotomous outcomes, these are presented as relative or absolute estimates (8,9). Understanding both is a core evidence-based practice (EBP) competency (2,10). Additionally, optimal dietetic practice should consider certainty of evidence, which indicates how much confidence clinicians and patients can place in an effect estimate (11).

Despite the importance of these competencies, little is known about whether Australian Accredited Practicing Dietitians’ (henceforth referred to as dietitians) understand effect estimates, an important competency in clinical decision-making. We therefore assessed their understanding of three absolute estimates (risk difference [RD]/absolute risk reduction [ARR], risk in treatment vs. risk in control groups and number needed to treat [NNT]) and two relative estimates (relative risk [RR]/relative risk reduction [RRR] and odds ratio [OR]/relative odds reduction [ROR]). We also assessed their confidence in determining the correct effect size, their perceived usefulness of each estimate and perceived usefulness of having certainty of evidence alongside effect estimates.

## METHODS

### Study design, sample and survey distribution

We conducted an anonymous web-based pilot survey using Qualtrics. Eligible participants were English-proficient dietitians involved in either clinical practice, teaching or research. The University of Wollongong Human Research Ethics Committee approved the protocol (Ethics Number: 2024/220). Participants provided tacit consent by completing the survey. We first distributed the survey through Dietitians Australia, then through the Nutrition Society of Australia, X (formerly Twitter), LinkedIn and personal networks. The survey remained open from October 1, 2024 to February 3, 2025. We used Qualtrics reCAPTCHA, bot detection and IP checks to prevent duplicate or automated responses.

### Questionnaire development

We adapted a questionnaire developed for physicians to assess understanding of effect estimates (12). To examine whether understanding differed by effect size, we created two survey versions: one with a large effect size (ARR = 6.0%) and one with a trivial effect size (ARR = 0.2%). Participants were randomly assigned to either version upon accessing the survey link.

Each version included two sections. The first collected demographic and background information; the second assessed dietitians’ understanding, confidence and perceived usefulness of effect estimates. The second section began with a clinical scenario based on a hypothetical SRMA of randomized controlled trials comparing five dietary interventions with usual diet for reducing myocardial infarction over 5 years (13). We asked participants to complete the survey without using search engines or generative AI.

For each effect estimate, participants selected one of four effect-size interpretations: trivial (probably not important), small (probably important), moderate (surely important), or large (very important) (14). Then using 7-point Likert-type scales (1 = not useful at all to 7 = extremely useful), they rated confidence in determining the correct effect size, the usefulness of each estimate and the usefulness of presenting certainty of evidence.

### Data analysis

We analyzed only fully completed surveys and excluded incomplete ones. We used descriptive statistics to summarize participant characteristics. For understanding, we reported correct responses as frequencies and percentages with 95% confidence intervals (CIs). For confidence, perceived usefulness and usefulness of certainty of evidence, we reported means, standard deviations (SDs) and 95% CIs. We used binomial tests to assess whether understanding differed from the expected proportion by chance (Microsoft Excel 97–2003).

## RESULTS

Approximately 7,697 dietitians were sent the survey link, 27 started it and were randomized to either the trivial (n=16) or large (n=11) effect-size version, and 13 fully completed it (trivial, n=8; large, n=5). All completers were female, 38.5% held a doctoral degree, 77% completed epidemiology or biostatistics courses and 46.2% completed EBP related courses (**Table 1**).

**Table 1.** Characteristics of completers, n = 13.

| Characteristic | Frequency (%) |
|---|---|
| **Sex** |  |
| Female | 13 (100.0%) |
| **Primary work setting** |  |
| Clinical Nutrition | 4 (30.8%) |
| Primarily Research | 4 (30.8%) |
| Both Research & Teaching | 2 (15.4%) |
| Industry | 1 (7.7%) |
| Private Practice | 1 (7.7%) |
| Public Health | 1 (7.7%) |
| **Years involved in research** |  |
| 1 to 5 years | 4 (30.8%) |
| 6 to 10 years | 0 (0.0%) |
| 11 to 15 years | 1 (7.7%) |
| 16 to 20 years | 1 (7.7%) |
| 21 years or more | 3 (23.1%) |
| None | 4 (30.8%) |
| **Clinical work arrangement** |  |
| Full time (≥ 35 hours/week) | 4 (30.8%) |
| Part time (< 35 hours/week) | 4 (30.8%) |
| Per diem (as needed) | 1 (7.7%) |
| I am not involved with clinical practice | 4 (30.8%) |
| **Years in clinical practice** |  |
| 1 to 5 years | 5 (38.5%) |
| 6 to 10 years | 2 (15.4%) |
| 11 to 15 years | 0 (0.0%) |
| 16 to 20 years | 2 (15.4%) |
| 21 years or more | 3 (23.1%) |
| None | 1 (7.7%) |
| **Highest degree in nutrition or dietetics** |  |
| Doctoral Degree | 5 (38.5%) |
| Bachelor's Degree | 3 (23.1%) |
| Master’s Degree | 3 (23.1%) |
| Doctoral Degree in Progress | 2 (15.4%) |
| **Training on epidemiology/biostatistics** |  |
| Yes | 10 (76.9%) |
| No | 3 (23.1%) |
| **Training on data presentation methods and certainty/strength of evidence** |  |
| Yes, both | 7 (53.8%) |
| Yes, data presentation methods | 2 (15.4%) |
| Yes, certainty/strength of evidence | 1 (7.7%) |
| No | 3 (23.1%) |
| **Training on EBP** |  |
| No | 7 (53.8%) |
| Yes | 6 (46.2%) |
| **Participated in EBP-focused CPD/seminar/workshop** |  |
| No | 10 (76.9%) |
| Yes | 3 (23.1%) |

Overall, correct understanding was higher for absolute estimates than relative ones (30% vs. 18.2%). Understanding was highest for risk in treatment vs. risk in control groups (41.7%) and lowest for RD/ARR, RR/RRR, and OR/ROR (all 18.2%). Confidence was highest for RR/RRR [Mean (SD): 4.18 (1.25)] and lowest for NNT [3.40 (1.17)]. Perceived usefulness was highest for OR/ROR [4.69 (1.18)] and lowest for NNT [3.75 (1.76)]. Having certainty of evidence was perceived as useful [5.69 (1.03)]. **Table 2** shows the understanding, confidence and usefulness of five effect estimates. Binomial tests suggest that understanding was not significantly higher than what would be expected by chance alone (p-values > 0.05, **Table 2**).

**Table 2.** Correct understanding, confidence and perceived usefulness (n=13)

| **Understanding** |  |  |  |  |
|---|---|---|---|---|
| **Approach** | **Total # of respondents** | **n (%) correct** | **95% CI** | **Binomial test\* (π > 25%)** |
| RR/RRR | 11 | 2/11 (18.2%) | [5.1%–47.7%] | p = 0.803 |
| OR/ROR | 11 | 2/11 (18.2%) | [5.1%–47.7%] | p = 0.803 |
| RD/ARR | 11 | 2/11 (18.2%) | [5.1%–47.7%] | p = 0.803 |
| NNT | 10 | 3/10 (30.0%) | [10.8%–60.3%] | p = 0.474 |
| Risk in both groups | 12 | 5/12 (41.7%) | [19.3%–68.0%] | p = 0.158 |
| **Confidence** |  |  |  |  |
| **Approach** | **Total # of respondents** | **Mean (SD)** | **95% CI** |  |
| RR/RRR | 11 | 4.18 (1.25) | [3.34–5.02] | - |
| OR/ROR | 11 | 3.55 (1.04) | [2.85–4.24] |  |
| RD/ARR | 11 | 3.55 (0.82) | [2.99–4.10] |  |
| NNT | 10 | 3.40 (1.17) | [2.56–4.24] |  |
| Risk in both groups | 12 | 3.83 (1.47) | [2.90–4.77] |  |
| **Perceived usefulness** |  |  |  |  |
| **Approach** | **Total # of respondents** | **Mean (SD)** | **95% CI** |  |
| RR/RRR | 13 | 4.46 (1.39) | [3.62–5.30] |  |
| OR/ROR | 13 | 4.69 (1.18) | [3.98–5.41] |  |
| **Understanding** |  |  |  |  |
|  |  |  |  | - |
| RD/ARR | 12 | 4.25 (1.42) | [3.35–5.15] |  |
| NNT | 12 | 3.75 (1.76) | [2.63–4.87] |  |
| Risk in both groups | 13 | 4.46 (1.76) | [3.40–5.53] |  |

CI = confidence interval
\* For all types of estimates, the percent correct did not statistically vary from 25% based on binomial testing.

## DISCUSSION

Dietitians showed better understanding of absolute estimates than relative ones, however, they reported highest confidence and perceived usefulness for relative estimates. This disconnect suggests dietitians may judge confidence and usefulness based on familiarity with relative estimates rather than the actual understanding (8,15).

This is the first survey to assess Australian dietitians’ understanding of treatment effect sizes and views on certainty of evidence for clinical decision-making. However, the small sample limits generalizability. In addition, the use of effect size category ‘trivial’ (14) may also have influenced responses, as dietitians may be unfamiliar with what magnitude should be interpreted as trivial rather than small (16-19). This limitation, along with understanding not differing from the expected proportion by chance, highlights the need for standardized EBP competency training on absolute and relative estimates, effect size classification and certainty of evidence.

Participants echoed this need in end-of-survey feedback, with one noting, “I am now unsure of my interpretation of effect sizes… I need more training!” Another suggested embedding EBP “into many facets of nutrition and dietetics learning and practice.”

Future research should include larger and more diverse samples, including dietetic students and interns, and assess whether understanding differs between dietitians in academic settings and clinical practice. Intervention studies should test whether targeted education improves these competencies.

## CONCLUSION

Preliminary results from a small dietetic sample showed better understanding of absolute effect estimates than relative ones. The disconnect between understanding, confidence and perceived usefulness suggests a need for training in effect size, and evaluation in larger sample.

## Data Availability

All data produced in the present work are contained in the manuscript.

